# Maternity care professionals’ preparedness for and experiences of screening and responding to disclosures of domestic violence in the peripartum period: a protocol for a qualitative evidence synthesis

**DOI:** 10.1101/2024.04.25.24306402

**Authors:** Laura O’Shea, Melissa Corbally, Déirdre Daly

## Abstract

**Background:** Maternity care professionals, such as midwives, public health nurses, doctors, and social workers, are in the unique position of having regular contact with women in the peripartum period. They are well-placed to recognise and respond to disclosures of domestic violence, however many lack confidence and feel unprepared for this in practice. While there are screening tools used for enquiry about domestic violence in pregnancy, there are variations in the tools used, the frequency and timing of enquiry, and the response/referral pathways across professions. Research exists on the role of health care professionals such as midwives, doctors, and nurses with regards to domestic violence, however little is understood about the collective and shared experience of maternity care professionals who screen for and respond to domestic violence in the peripartum period.

**Methods:** A qualitative evidence synthesis of maternity care professionals’ preparedness for and experiences of screening and responding to disclosures of domestic violence in the peripartum period will be conducted. Qualitative studies of any design, and mixed method and other design studies where qualitative data can be extracted will be considered for inclusion. A systematic search of the following electronic bibliographic databases will be conducted: ASSIA, CINAHL, EMBASE, Maternity and Infant Care, MEDLINE, PsycINFO and SocINDEX. The Critical Skills Appraisal Programme (CASP) qualitative studies tool will be used to assess methodological quality of included studies. Data synthesis will involve three sequential stages, coding, development of descriptive themes and generation of analytical themes. Confidence in findings will be assessed using the Grading of Recommendations Assessment, Development and Evaluation-Confidence in the Evidence from Reviews of Qualitative research (GRADE-CERQual) tool.

**Discussion:** This QES will provide a deeper understanding of maternity care professionals’ preparedness for and experiences of screening and responding to domestic violence disclosures in the peripartum period. The findings will, potentially, identify what aspects of education and preparedness work well and what might be improved.

## Introduction

Domestic violence, also called intimate partner violence (IPV) or domestic abuse, refers to physical, emotional, sexual, and financial abuse within close adult relationships, most often intimate partner relationships. It also encompasses abuse in the form of coercive and controlling behaviours [1]. Domestic violence is addressed in international and national legislation to varying degrees. The Council of Europe Convention on preventing and combating violence against women and domestic violence is the first European instrument that aims to legally prevent or legislate for the prevention of gender-based violence. It aims to protect victims of violence and punish perpetrators of domestic violence. It provides a framework of measures to prevent and combat domestic violence at international and national levels. These include raising awareness, data collection, legislation, training, education, and improvement of resources. As of January 2024, the Convention has been signed by all EU Member States and ratified by 22 states [2]. In Ireland, domestic violence is dealt with in both criminal and civil law. The first significant legislation in Ireland regarding domestic violence was included in the Family Law (Maintenance of Spouses and Children) Act 1976 which provided statutory entitlement to apply for a barring order. Following this, the Family Law (Protection of Spouses and Children) Act 1981 was introduced and provided for protection orders. In 1996, the Domestic Violence Act introduced the safety order which was effectively a more permanent protection order. In more recent years, The Domestic Violence Act 2018 improved domestic violence legislation in Ireland, consolidated existing law on domestic violence and provided additional protections. This legislation also introduced two new criminal offences: forced marriage and coercive control [3].

A European Union Agency for Fundamental Rights (FRA) study found that just over one in five women experienced domestic violence in the European Union [4]. Globally, almost one in three women are subjected to intimate partner violence at least once in their lives [5]. The lockdown restrictions during the Covid-19 pandemic further compounded this issue, leaving women more isolated and made it more difficult to seek assistance and access pathways to safety [6]. Pregnancy and the postpartum period are identified as times of increased risk of domestic violence as this is often when violence first occurs or escalates [1]. A systematic review and meta-analysis on the worldwide prevalence of domestic violence in pregnancy found that there is variance in the rates of domestic violence reported [7]. The review found that this is due to the use of various definitions of domestic violence, the measurement strategy, and the socio-cultural context of the population studied [7]. The authors found that the global prevalence rate of domestic violence in pregnancy was 25.0% (95% CI 20.4–30.7). Prevalence figures were higher in Africa (36.1%, 95% CI 27.7–45.4) than in Asia (32.1%, 95% CI 22.7–43.2), and in South America (25.6%, 95% CI 21.1–30.7) and North America (20.4%, 95% CI 6.9–47.1). The lowest prevalence was reported for Europe (5.1%, 95% CI 3.4–7.5) [7]. This review retrieved data from over 50 countries and found that overall, one in ten mothers were exposed to physical violence, one in five to psychological violence and one in twenty to sexual violence [7].

In 2021, in Ireland, a total of 152 women reported to Women’s Aid that they had been abused in pregnancy, while 41 had suffered a miscarriage due to domestic violence [8]. Domestic violence in pregnancy may lead to mortality and/or lifelong multiple morbidities for both the woman and fetus; these include but are not limited to, first and second trimester miscarriage, placental abruption, preterm labour, low birth weight infant, depression, and anxiety [9].

Women’s increased contact with healthcare professionals during pregnancy offers a unique opportunity to identify and support those who are experiencing domestic violence. Research demonstrates that a protocol of routinely asking all women if they are experiencing domestic violence in pregnancy supports women to disclose and seek supports [10, 11, 12]. Screening aims to identify women who have experienced or are currently experiencing domestic violence and to offer interventions [13]. The World Health Organisation (WHO) recommends that healthcare professionals enquire about domestic violence in the peripartum period [14]. Whilst a Cochrane review found insufficient evidence to recommend asking all women about domestic violence in healthcare settings, it did state that screening may be beneficial and that healthcare professionals should be trained to ask women who show signs of abuse or those in high-risk groups and provide them with a supportive response and information [13]. There are many validated screening tools used globally, the most common tools tested are the Abuse Assessment Screen, Woman Abuse Screening Tool, and the HITS (Hurts, Insults, Threaten, Scream) tool [15]. However, there is a lack of consensus regarding the types of tools used, the frequency and timing of enquiry, and the response/referral pathways across healthcare professions [16,17].

Midwives, public health nurses, doctors, and medical social workers are in the unique position of having regular contact with women during pregnancy, labour and birth and the postnatal period. Research has highlighted a lack of consistent domestic violence education with many healthcare professionals lacking confidence and feeling unprepared to screen for and respond to disclosures of violence [16, 18, 19, 20, 21]. Previous research in this area has mainly focused on barriers to women disclosing domestic violence [15, 22]. While there is some research regarding the role and experiences of health care professionals such as midwives, doctors and nurses [18,20] little is understood about the collective experience of maternity care professionals who screen for and respond to domestic violence in the peripartum period. This qualitative evidence synthesis (QES) aims to integrate the findings from studies that explore the experiences of the relevant maternity healthcare professionals and identify what enables them to feel more prepared to enquire about and respond to women’s disclosures of domestic violence. The findings of this QES will deepen understanding of the collective experiences of maternity care providers in this situation and aid in identifying potential gaps in clinical practice and education around this important issue.

## Protocol

### Inclusion criteria

The SPIDER (sample, phenomenon of interest, design, evaluation, and research type) tool is used to develop the key concepts and the inclusion criteria in this QES.

**S**ample: Maternity health care professionals who provide peripartum care. These include midwives, obstetric nurses, public health nurses, community nurses, doctors, general practitioners, obstetricians, medical social workers/social workers.

**P**henomenon of **I**nterest: Maternity care professionals’ experiences of screening for and responding to disclosures of domestic violence in the peripartum period.

**D**esign: Qualitative studies of any design will be included. Mixed method design studies where qualitative data can be extracted and any studies that provide qualitative data, such as responses to open-ended questions in surveys, will also be considered for inclusion.

**E**valuation: Analytical themes that depict maternity health care professionals’ experience of screening for and responding to disclosures of domestic violence in the peripartum period.

**R**esearch type: English language, published, peer reviewed qualitative studies, and mixed methods or quantitative studies where qualitative data can be extracted separately will be considered for inclusion.

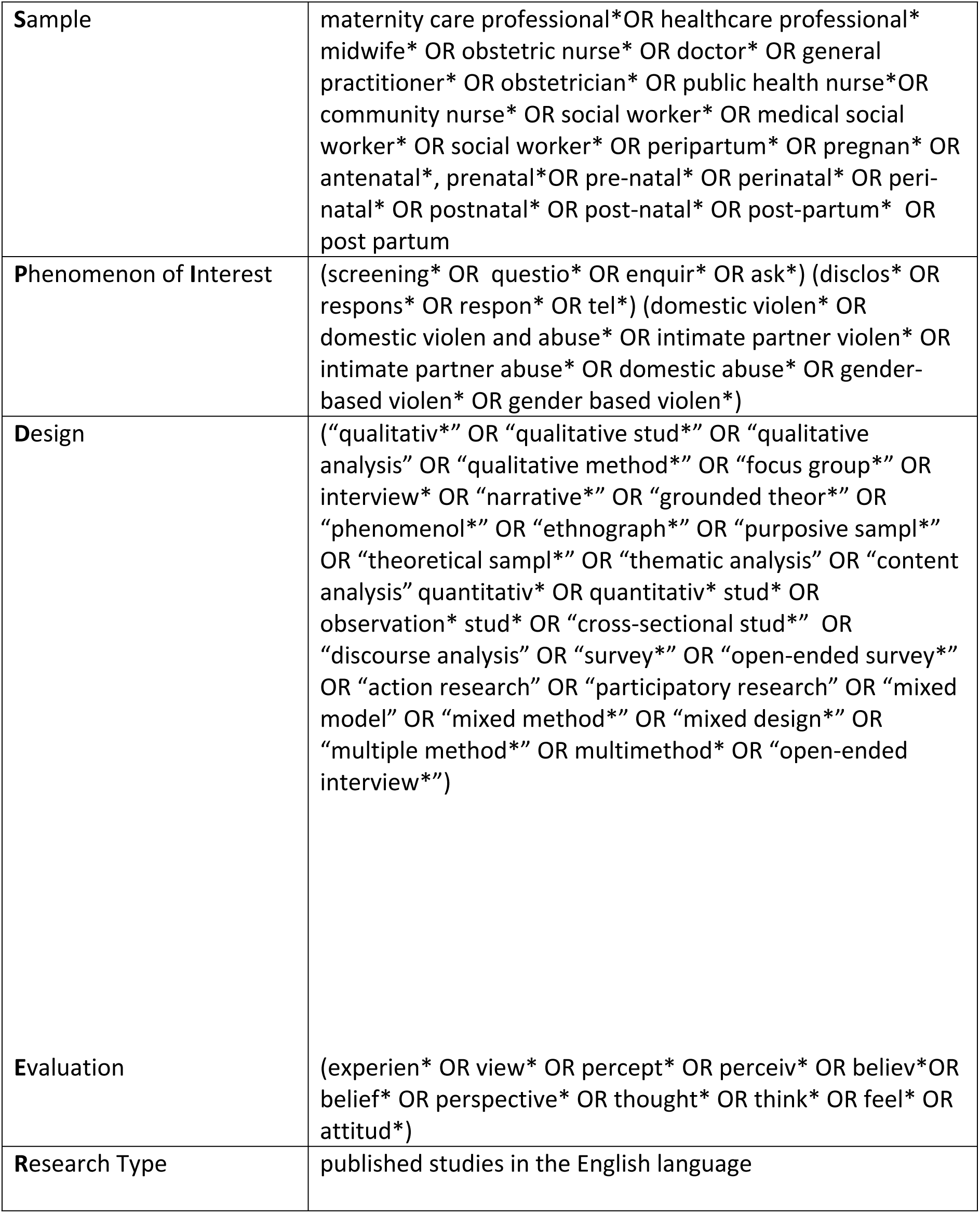

### Search strategy and study selection

Key guidance on the search strategy and inclusion process for QES details seven steps, the “7 S” approach [23]. This approach prompts the reviewer to consider the following- sampling of papers, sources, structured questions, search procedures, search strategies and methodological filters, supplementary searches, and strategies for reporting searches. An initial scoping search of CINAHL, MEDLINE and EMBASE was conducted to identify relevant studies for inclusion and further develop appropriate search terms. Search terms have been developed using the SPIDER acronym and will be adapted for each database searched (Table 1).

The following subject-appropriate bibliographic databases will be systematically searched for relevant studies: ASSIA, CINAHL, EMBASE, Maternity and Infant Care, MEDLINE, PsycINFO and SocINDEX. Several supplementary strategies to retrieve all relevant research will be conducted. These include reference checking of included studies, handsearching of journals and, when appropriate, contact with subject experts and authors in the field of domestic violence. Databases such as OpenGrey, GreyLit and GreyNet will also be searched to retrieve relevant grey literature. There will be no date restrictions applied during searches in order to include any seminal pieces of research. There will be no restrictions applied on geographic location or country of origin, however only English language articles will be included as we will not have access to translation services.

The retrieved citations will be exported to Endnote (version EN21) and duplicates will be removed. The remaining citations will be uploaded to Covidence, a screening and data extraction software tool to screen for eligibility. Twenty percent of selected studies’ titles and abstracts will be independently reviewed by two reviewers to assess if they meet the inclusion

criteria [24]. This step will aid in ensuring that there is an agreed and shared understanding of the criteria among reviewers. Once an agreement has been reached on the application of the inclusion criteria, screening of the remaining citations will take place. Screening will be a two-stage process including title and abstract screening and full text screening and review. Title and abstracts will be screened by two independent reviewers against the inclusion criteria. Following this, the full text of potentially relevant studies will be reviewed. Any disagreements that arise will be resolved through discussion until consensus is achieved or through an additional review by a third reviewer. The reasons for exclusion of studies reviewed at full text will be recorded in the final QES. The results of the search and the screening process will be reported in the final QES and presented in a Preferred Reporting Items for Systematic Reviews and Meta-analyses (PRISMA) flow diagram [25].

### Assessment of methodological quality

The assessment of methodological quality of included studies and the use of appraisal tools in QES is becoming increasingly popular [26]. Assessment of methodological quality of studies included in a QES can provide useful information on the robustness of the conduct of studies and the credibility of conclusions and increase understanding of the transferability of the findings [27]. The decision of which appraisal tool to use depends on the objectives of the QES, time, resources, and the expertise of the researcher [28]. The Critical Skills Appraisal Programme (CASP, 2018) qualitative studies tool, which uses 10 questions, will be used to assess methodological quality of included studies (Table 2). Two reviewers will independently assess all included studies. Consensus will be reached following discussion between the reviewers or assessment by a third reviewer, if required. All screened studies will undergo data extraction and synthesis as the findings from all included studies, irrespective of their methodological quality, may be applicable to the aim of the review.

**Table 2 Criteria for methodological quality assessment (CASP, 2018)**
|  |  |
| --- | --- |
| 1. Was there a clear statement of the aims of the research? | Yes/No/Can't Tell |
| 2. Is a qualitative methodology appropriate? | Yes/No/Can't Tell |
| 3. Was the research design appropriate to address the aims of the research? | Yes/No/Can't Tell |
| 4. Was the recruitment strategy appropriate to the aims of the research? | Yes/No/Can't Tell |
| 5. Was the data collected in a way that addressed the research issue? | Yes/No/Can't Tell |
| 6. Has the relationship between researcher and participants been adequately considered? | Yes/No/Can't Tell |
| 7. Have ethical issues been taken into consideration? | Yes/No/Can't Tell |
| 8. Was the data analysis sufficiently rigorous? | Yes/No/Can't Tell |
| 9. Is there a clear statement of findings? | Yes/No/Can't Tell |
| 10. How valuable is the research? | Yes/No/Can't Tell |

### Data extraction and synthesis

A purposively designed data extraction form will be created. This form will include information regarding the population studied, the characteristics of the study, the study context, the methods, and methodology used, details regarding recruitment, data collection methods and details regarding data analysis. Data extraction will be a two-stage process. First, the contextual data will be extracted. The second stage of data extraction involves extraction of the findings from each individual study, these will be organised systematically to enable data synthesis and analysis [29]. The thematic synthesis approach developed by Thomas and Harden will be used for data synthesis [30]. The synthesis will be performed in three sequential stages that may overlap to some degree: 1) line by line coding of data from primary studies, 2) development of descriptive themes, and 3) generating analytical themes. The first stage, coding, will be conducted independently by two review authors. These codes will be grouped into descriptive themes and following discussion within the review team analytical themes will be identified.

### Assessing confidence in the findings

It is important to assess both the quality of individual studies and the level of confidence in the evidence generated from the review of qualitative research. The QES findings will be assessed using the Grading of Recommendations Assessment, Development and Evaluation- Confidence in the Evidence from Reviews of Qualitative research (GRADE-CERQual) tool) [23]. GRADE-CERQual is based on assessment of four components: methodological limitations, coherence, adequacy of data, and relevance, and overall assessment of each component will be classified as high, moderate, low or very low as per the CERQual tool [23].

### Study status

The protocol has been registered on PROSPERO CRD42023464776. Initial scoping searches have been conducted. The proposed date for completion is January 2025.

### Ethical Approval

As this is a protocol for a qualitative evidence synthesis, ethical approval is not applicable.

## Discussion

Research has shown that most women are in favour of routine questioning if asked in a sensitive manner and by a well-trained health professional. It has been argued that such an approach leads to increased rates of disclosure [31, 32]. Previous research has mainly focused on barriers to women disclosing domestic violence. These barriers include shame, stigma, fear of judgement or not being believed and confidentiality concerns [13,22]. There is a lack of exploration of the collective experience of maternity healthcare professionals who screen for and respond to domestic violence, particularly their education and preparedness for this in clinical practice. Despite the evidence to suggest that screening is useful in identifying domestic violence [13] it appears that healthcare professionals may not always feel prepared to ask and respond to or respond effectively to a disclosure [33,34]. For example, in two studies conducted with healthcare professionals, doctors and nurses reported receiving little or no training in responding to domestic violence [35,36]. Research has found how challenging it can be for a healthcare professional to ask women about a history of domestic violence, either routinely or if a risk is identified [11]. Healthcare professionals identified a lack of knowledge, education and feeling unsupported, unprepared, and unconfident when screening for domestic violence as significant challenges in clinical practice. Previous studies also identified a lack of implementation of consistent guidelines, and lack of knowledge of the services or referrals that were available as barriers to healthcare professionals screening for domestic violence (16,37,38] Henriksen et al in their study, highlighted a lack of motivation by health professionals to screen women, due to a feeling of insecurity regarding what to do if violence was disclosed [39].

To our knowledge, this QES will be the first to synthesise qualitative data on the collective experiences and preparedness of maternity healthcare professionals in screening for and responding to domestic violence in the peripartum period. This QES aims to focus on what enables maternity healthcare professionals to feel prepared to enquire and respond to disclosures of domestic violence and to identify any professional discipline specific educational and clinical requirements.

## Data Availability

Deidentified research data will be made publicly available when the QES is completed and published.

https://osf.io/7ajvf

## Data availability Underlying data

No data are associated with this article

Registered on PROSPERO International prospective register of systematic reviews on 30/9/23

Available from https://www.crd.york.ac.uk/prospero/display_record.php?ID=CRD42023464776

## Reporting guidelines

Open Science Framework: Prisma-P checklist for Maternity care professionals’ preparedness for and experiences of screening and responding to disclosures of domestic violence in the peripartum period: a protocol for a qualitative evidence synthesis.

## Reflexivity statement

There is increasing recognition that reflexivity is an essential component of qualitative research [40]. Reflexivity is a process of continuous, collaborative practices that researchers adopt to consciously critique, appraise, and evaluate how their background or subjectivity may influence the research process [40]. This QES will utilise a reflective approach throughout the research process. To enhance the trustworthiness of the findings, a research diary will be kept documenting the process, reviewers will have frequent discussions regarding descriptive and analytical themes and quotations from primary studies will be included to support the findings of the review [41].

## Authors’ Contributions

LOS will undertake this QES as part of her PhD project. LOS conceptualised the review question under supervision of DD and MC. LOS, DD and MC had active roles in contributing to the design of the QES protocol. LOS drafted the protocol manuscript. DD and MC contributed to the writing through review and editing.

